# Move by move towards mental health: A pilot study on chess as a therapeutic approach in adolescents with mental disorders

**DOI:** 10.1101/2025.10.29.25338524

**Authors:** Sarah Gerhardt, Stefanie Hoier, Alexandra Seeger, Roland Schmidt, Larissa Weber, Konstantin Mechler, Tobias Banaschewski, Alexander Häge, Sabine Vollstädt-Klein

## Abstract

**Background:** Mental disorders affect approximately 14% of adolescents worldwide, often leading to persistent cognitive and emotional difficulties and reduced health-related quality of life (HRQoL). Executive functions (EF)—including cognitive flexibility, inhibition, attention, and working memory— are particularly impaired in many psychiatric conditions. Chess has recently been proposed as a low-cost cognitive remediation training (CRT). This pilot study investigated whether chess-based CRT could enhance EF and HRQoL in adolescents with psychiatric disorders.

**Methods:** A quasi-experimental design was conducted at the Department of Child and Adolescent Psychiatry, Mannheim, Germany (September 2022–April 2024). Participants aged 13–17 years were assigned to either a six-week chess intervention (experimental group, EG) or treatment as usual (control group, CG). Both groups received standard multidisciplinary therapy, while the EG additionally participated in weekly 90-minute chess sessions based on The King’s Plan for Kids. Cognitive flexibility (DCCS), inhibitory control (Stop-Signal Task), sustained attention (d2-R), and working memory (n-back task) were assessed alongside HRQoL (KIDSCREEN-27). Data were analyzed using t-tests.

**Results:** Thirty-three adolescents were included (19 EG, 14 CG; 82% female). While no significant group differences emerged for cognitive flexibility, inhibitory control, or sustained attention, the EG showed significantly faster reaction times in the working memory task (p = .016, d = 0.79), suggesting improved cognitive efficiency. Psychological well-being increased significantly in the EG compared to the CG (p = .035, d = 0.67), whereas physical well-being showed a non-significant upward trend.

**Conclusion:** Chess-based CRT was associated with enhanced psychological well-being and improved working memory efficiency in adolescents with psychiatric disorders. Although other EF measures did not show significant changes, findings support the feasibility and potential clinical value of chess as an engaging, low-risk adjunct to standard therapy. Larger randomized trials are needed to confirm these preliminary results.

## INTRODUCTION

According to the World Health Organization (World Health Organisation WHO, 2021) 14% of 10-to 19-year-olds worldwide experience mental health problems, most commonly emotional and behavioral disorders. The associated costs pose significant challenges for both healthcare systems and society (European Union OECD, 2016). This is particularly concerning as, in some cases, these disorders persist into adulthood—around half of adults with mental disorders report symptom onset during childhood or adolescence—increasing the risk of long-term impacts on development, quality of life, and daily functioning (Fuchs et al., 2012; Hammud et al., 2023; Klipker et al., 2018). Mental disorders in children and adolescents, such as attention deficit hyperactivity disorder (ADHD), anorexia nervosa, or depression, are often linked to cognitive impairments, particularly in executive functioning (EF), including cognitive flexibility, inhibition, sustained attention, and working memory (e.g., Cortese et al., 2015; Goodall et al., 2018; Lima et al., 2018; Tchanturia et al., 2017; X. Wang et al., 2023). These children and adolescents also typically report a significantly lower quality of life (QoL), specifically health-related QoL (HRQoL), than their peers—often comparable to or worse than that of individuals with physical illnesses (Bastiaansen et al., 2020; Dey et al., 2012; Klipker et al., 2018; Otto et al., 2021; Sawyer et al., 2002). Furthermore, difficulties in EF and poor self-regulation have been suggested to negatively impact QoL in children with emotion regulation disorders, indicating a possibly causal relationship (Hammud et al., 2023).

Conversely, better cognitive functioning is not only associated with higher QoL (Hammud et al., 2023; X. Wang et al., 2023), but also with reduced risk of disease recurrence (Y. Wang et al., 2022) and greater therapy engagement or shorter hospital stays (Jeste et al., 2003; McKee et al., 1997). EF can be improved throughout childhood and adolescence through training and targeted interventions (Diamond, 2013; Serpell & Esposito, 2016; Tchanturia et al., 2017). Effective therapeutic interventions that target EF are therefore essential for children and adolescents with mental disorders. To this end, cognitive training can enhance psychotherapeutic outcomes (Bowie & Harvey, 2006). Diamond (2013) further emphasizes the importance of supporting children in developing strong EF, given its ties to QoL. Specifically, Cognitive Remediation Training (CRT) has been shown to improve cognitive functioning and is increasingly used in this context (Cella et al., 2016). It has shown promising effects on inattention symptoms and working memory in ADHD (Cortese et al., 2015), cognitive flexibility, problem-solving, and QoL in ASD (Hajri et al., 2019; Pasqualotto et al., 2021), as well as EF and attention in psychosis (Anagnostopoulou et al., 2019). However, mixed results have also been reported—for example, in anorexia nervosa (Tchanturia et al., 2017). Similarly, chess has gained popularity as a cognitive and social training tool. Reflecting its global appeal and potential educational benefits, the European Parliament (Declaration of the European Parliament of 15 March 2012 on the introduction of the programme ‘Chess in School’ in the educational systems of the European Union., 2012) recommended the inclusion of chess in school curricular, as play—including chess—has been shown to support both cognitive and social development (Aciego et al., 2012; Cordier et al., 2009). Playing chess engages multiple cognitive functions, such as cognitive flexibility, inhibition, attention, memory, problem-solving, and planning (Bilalić et al., 2009; Unterrainer et al., 2006). Chess players in grades 5 to 9 have demonstrated better mathematical problem-solving and metacognitive abilities compared to control groups (Kazemi et al., 2012). Similarly, children who play chess show improvements not only in cognitive abilities such as EF (Ramos et al., 2018) but also in socio-emotional aspects such as self-confidence (Aciego et al., 2012).

Despite growing interest, research on the specific cognitive effects of chess in young people with mental disorders remains limited—especially regarding EF and QoL. Studies, primarily focusing on ADHD, have reported positive effects of chess on concentration, listening skills, and task focus in children aged 11 to 13 (Mohammad Nour ElDaou & El-Shamieh, 2015) as well as reduced ADHD symptoms—including inattention and hyperactivity—in children aged 6 to 17 (Blasco-Fontecilla et al., 2016). More broadly, research on chess’s effects on cognitive functions in children and adolescents is mixed and typically yields small to moderate effect sizes (Sala & Gobet, 2016).

Regarding QoL, a study in school children examined important aspects of QoL by self-report measures of school satisfaction, engagement as well as self-efficacy, and found self-perceived positive effects by chess at all ages (Chitiyo et al., 2023). Elementary school children showed highest effects, especially related to self-efficacy and engagement in school learning. In an own pilot study in adults, we found that female chess experts showed increased satisfaction with life and less physical complaints than the average female population (Vollstädt-Klein et al., 2010). A study in older adults found that a 12-week chess-training protocol with two 60-minute sessions per week improved cognition (especially related to attention, processing speed and executive function) and QoL (Cibeira et al., 2021).

Moreover, existing findings suggest that chess has potential as an effective therapeutic intervention for enhancing cognitive functioning and QoL also in the population of adolescents. It also offers several advantages: it is simple, cost-effective, free from side effects, and generally well-accepted by children and parents. Furthermore, chess can be implemented in various settings (inpatient, outpatient, or at home) and promotes social integration, making it a versatile intervention (Agarwal, 2023; Blasco-Fontecilla et al., 2016).

Consequently, an innovative therapeutic approach for children and adolescents with mental disorders involves chess-based CRT. This pilot study aimed to explore whether chess-based CRT could improve cognitive flexibility, inhibitory control, sustained attention, and health-related quality of life (HRQoL—both physical and psychological well-being) in a naturalistic sample of adolescents with various mental disorders receiving inpatient psychiatric and psychotherapeutic treatment. We hypothesized that chess training, delivered as a six-week add-on intervention, would improve (1) cognitive flexibility, (2) inhibitory control, (3) sustained attention, (4) working memory and both (5) physical and (6) psychological well-being compared to treatment as usual (TAU) alone.

## MATERIALS AND METHODS

### Procedure and participants

This quasi-experimental pilot study was conducted at the Department of Child and Adolescent Psychiatry and Psychotherapy (CAP), Central Institute of Mental Health (CIMH), Mannheim, Germany between September 2022 and April 2024. The non-equivalent group design resulted in a 2 (group: experimental, control) x 2 (time: pre-test, post-test) mixed-factor design, with time as the within-subjects factor and group the between-subjects factor. Group allocation was quasi-randomized to either the experimental group (EG; enrolment and data collection September 2022 until September 2023) or the control group (CG; data collection and enrolment July 2024 until April 2024). An a priori power analysis for one-sided t-tests for independent samples was conducted out using G*Power 3.1.9.7 (Faul et al., 2009). A medium effect size of d = 0.5 (Cohen, 1988) was targeted. Based on a desired power of (1 - β) = .80, an error probability of α = .05 and groups of equal size, a total sample size of 102 adolescents was targeted, with 51 participants in each EG and CG.

Participants between the ages of 13 and 17 years with sufficient verbal and written communication abilities were recruited at the CAP. To ensure full participation over the six-week study period, patients (in-house, outpatient, or in inpatient-equivalent settings) were included during the first three weeks after treatment initiation, provided their treatment duration was expected to be adequate. Ongoing medication use (e.g., antidepressants, ADHD medication) was permitted during study participation.

Exclusion criteria included severe neurological, psychiatric, or internal comorbidities, which in the opinion of the treating physicians, contraindicates participation in the study, a body mass index (BMI) below 14, and placement under the German Civil Code (BGB). As financial compensation, participants received €20 after completing all assessments, which was donated by the Munich Chess Foundation (Münchener Schachstiftung, https://schachstiftung-muenchen.de). The overall study of which this data derived from was pre-registered at ClinicalTrials.gov (NCT01401491). Study procedures were in line with the recommendations of the World Medical Association (revised Declaration of Helsinki) as well as in accordance with good clinical practice guidelines, and were approved by the local Ethics Committee of the Medical Faculty Mannheim, Heidelberg University (approval # 2021-593).

After identifying eligible patients using the clinic’s patient management system, study participation was proposed by their practitioner. Written informed consent was obtained from the legal guardians and informed assent was obtained from the children and adolescents. All parties were informed via telephone, email, or in person and received written information about the study.

Data were collected at two time points, see Figure 1, using the electronic data capture tool REDCap, hosted locally at our institute (Harris et al., 2019): before (A1) and after (A2) the six-week study period. Each assessment lasted approximately two hours and included several neuropsychological tasks and questionnaires, administered in the same order and at approximately the same time of day for all participants.

**Figure 1:**
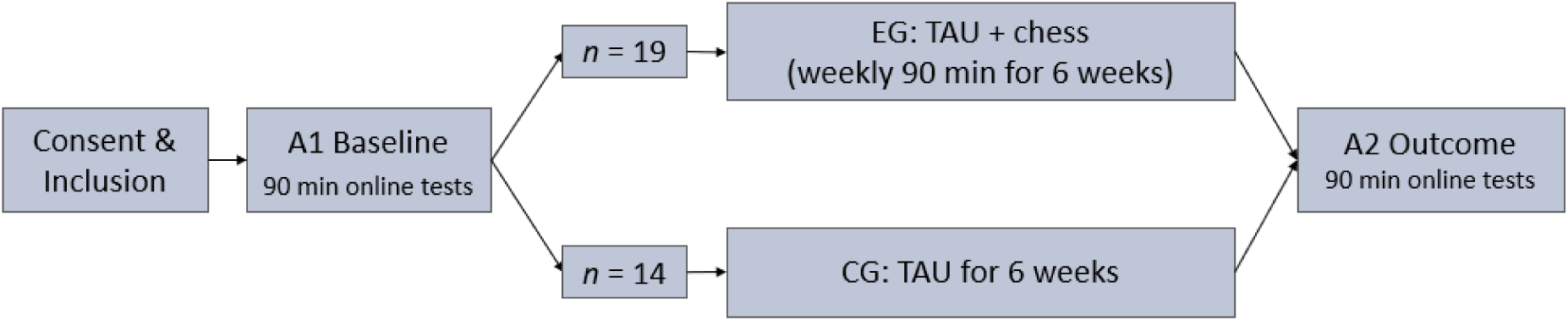
Study procedure including two assessment time points, A1 and A2. EG =experimental group, CG = control group, TAU = treatment as usual.

Both the control group (CG) and experimental group (EG) received treatment as usual, delivered by a multidisciplinary team. This included psychotherapy—primarily cognitive behavioral therapy (CBT)— as a core component, along with pharmacotherapy when necessary.

### Chess training

Participants in the EG took part in a 90-minute, in-person chess training led weekly in the late morning by a chess trainer (RS and AS) over the six-week study period. First, the basic rules of chess were introduced using a demonstration board and video assistance based on the established training concept Smart by Chess – The King’s Plan for Kids (Kindermann & Exler, 2023). This concept teaches the thinking and problem-solving strategies of chess players in an accessible way, aiming to develop fundamental cognitive skills through an engaging, creative, and playful approach. Each session began with chess yoga for relaxation and concentration. The trainer then introduced theoretical concepts and integrated physical exercises, role-playing, and concentration techniques. This was followed by active chess play, supported with guidance and tips. Sessions were balanced between theory and practice. For this study, the trainer (RS) adapted The King’s Plan for Kids to better suit the adolescent age group.

### Psychometric questionnaires

The self-report KIDSCREEN-27 questionnaire was used to assess HRQoL in children and adolescents aged 8 to 18 years (Ravens-Sieberer et al., 2014). It consists of 27 items across five scales: Physical well-being, Psychological well-being, Autonomy and Parent relations, Social support and peers, and School environment. Internal consistency is good to excellent (α = .80–.84), and test-retest reliability over two weeks ranges from .61 (social support and peers) to .74 (school environment). For this study, the Physical well-being scale (assessing activity level, energy, and health—lower scores reflect exhaustion, low energy, and feeling unwell) and the Psychological well-being scale (covering mood, emotions, self-perception, and life satisfaction—lower scores indicate low self-esteem, unhappiness, and depressive feelings) were used.

### Neuropsychological tasks

All participants completed neuropsychological tasks (35 minutes) and, after a short break, questionnaires (30 minutes). Tasks were administered via an in-house JavaScript-based platform. For the present analyses, the Dimensional Change Card Sort Test (DCCS; (Zelazo, 2006)), the Stop-Signal Task (SST;(Sebastian et al., 2013)), the d2 Test of Attention – Revised (d2-R; (Brickenkamp et al., 2010)), and the n-back working memory task (Callicott et al., 1999) were used to assess cognitive flexibility, inhibitory control, attention/concentration, and working memory capacity respectively.

DCCS performance was scored using the two-vector method (Zelazo et al., 2014), which combines accuracy and reaction time (RT) for participants with >80% accuracy. This method captures the RT “cost” associated with EF (Davidson et al., 2006). Each vector ranges from 0 to 5, yielding a total score from 0 to 10. For participants with ≤80% accuracy, the score equaled the accuracy vector.

Higher scores indicate better performance. The change in DCCS scores from pre- (A1) to post-test (A2) was used; positive differences reflect improvement.

Inhibitory control was assessed using the Stop-Signal Reaction Time (SSRT), calculated via the race model (Logan & Cowan, 1984) by subtracting the mean Stop Signal Delay (SSD) from the mean RT in correct go trials. Shorter SSRTs reflect better inhibitory control. Negative SSRT differences (A2 minus A1) indicate improvement, with greater negative values reflecting stronger gains.

For the d2-R, sustained attention (KL) was calculated by subtracting missed targets (AF) and confusion errors (VF) from the total correct targets (BZO): KL = BZO – AF – VF. Higher KL values indicate better attention. Processing speed (BZO) and accuracy (F%) were also derived, with F% = 100 × (AF + VF) / BZO; lower F% indicates higher accuracy. Only data from trials 2–13 were used (excluding first and last lines). Change scores between A2 and A1 were used, with positive values indicating improvement.

Working memory capacity was assessed using the n-back working memory task in a low (0-back) and high cognitive load condition (2-back). Outcome measures include the error rate (in %) as well as reaction times in the 2-back condition.

### Statistical analyses

Statistical analyses were conducted using IBM SPSS Statistics version 29.0.0 (IBM Corp., 2022) with a significance level of α = .05. Group comparisons between the EG and CG were performed for the within-group mean change in the respective variable between times A2 and A1 as dependent variables. Two-tailed t-tests for independent samples were used for interval-scaled variables, while χ²-tests were applied to categorical and dichotomous variables. One-tailed t-tests with α = .05 were used to test the directional hypotheses, accompanied by 90% bias-corrected and accelerated (BCa) confidence intervals. To address potential violations of assumptions—particularly normality and homogeneity of variance—bootstrapping with 10,000 iterations was used to compute confidence intervals and conduct inferential tests. Dependent variables included the change in DCCS performance score (cognitive flexibility), SSRT and SSD (inhibitory control), d2-R KL score (sustained attention), error rate and reaction times for correct responses (working memory), and the sum scores for the Physical well-being and Psychological well-being scales. As reaction times are typically measured on a ratio scale, changes in reaction times were calculated as percentage related to A1 reaction times. For all other variables (interval scales) mean differences between A1 and A2 were used.

## RESULTS

### Sample, sociodemographics, and comorbidities

After pre-screening, 92 patients in the CAP met the criteria for potential participation, out of which 33 could be included in the study (19 EG, 14 CG; see CONSORT Flow Diagram, Appendix E). Analyses included only participants with complete data for the relevant neuropsychological tests and questionnaires at both A1 and A2. Some participants were excluded from specific analyses due to invalid or missing data. According to Verbruggen et al. (2019), SSRT scores were invalid for 12 participants (9 EG, 3 CG) (see Section 2.5.1.2) and were excluded. Two EG participants were excluded from the d2-R task analyses due to missing A1 data but had complete data for all other measures. Additionally, four participants (1 EG, 3 CG) were excluded from the physical well-being analyses, and one (CG) from the psychological well-being analyses, due to missing data at A1 or A2.

The sample comprised 82% girls and 18% boys, with 15 girls and 4 boys in the EG, and 12 girls and 2 boys in the CG. This refers to biological sex; gender identity was not recorded. Participants in both groups were between 13 and 17 years old (M = 15.24, SD = 1.23), with a mean age of M = 15.16 (SD = 0.96) in the EG and M = 15.36 (SD = 1.55) in the CG. For main diagnoses, see Appendix A1. Each participant in the EG attended at least two chess training sessions, with a maximum of seven sessions (M = 4.68, SD = 1.73).

### Neuropsychological data

DCCS performance scores were generally high, indicating good *cognitive flexibility*, with participants achieving over 80% of the maximum score at A1. The EG showed less improvement from A1 to A2 compared to the CG; however, the difference was not significant (t(16.57) = −1.33, p = .101, d = −0.52, ΔM = −0.67, BCaCI₉₀% = [−1.58, 0.11]).

The SSRT in the EG decreased from A1 to A2, indicating an improvement in *inhibitory control*. In contrast, the CG showed a decline, with an increase in mean SSRT (M = 4.94, SD = 96.84). A one-tailed t-test for independent samples revealed a non-significant effect (t(19) = −1.02, p = .161, d = −0.45, ΔM = −34.58, BCaCI₉₀% = [−96.49, 22.45]). Similarly, the EG showed an increase in mean SSD (M = 49.53, SD = 79.83), while the CG demonstrated a slight decrease (M = −10.94, SD = 123.37). However, group differences were not significant (t(19) = 1.32, p = .102, d = 0.58, ΔM = 60.47, BCaCI₉₀% = [−2.67, 130.41]).

The EG showed a greater but non-significant improvement in *sustained attention* (d2-R KL) from A1 to A2 (M = 18.88, SD = 29.29) than the CG (M = 18.50, SD = 13.73; t(29) = 0.05, p = .482, d = 0.02, ΔM = 0.38, BCaCI₉₀% = [−12.17, 12.55]). Likewise, neither BZO nor F% yielded significant results (BZO: t(29) = 0.67, p = .253, d = 0.24, ΔM = 12.13, BCaCI₉₀% = [−14.17, 39.46]; F%: t(29) = 0.43, p = .335, d = 0.16, ΔM = 2.55, BCaCI₉₀% = [−6.58, 12.04]).

The performance in the *working memory task* - measured by the number of errors in the N-Back task in the high cognitive load condition (2-back) - did not change from A1 to A2, neither in the EG (t(18) = .925, p = .367) nor in the CG (t(13)=.448, p = .663). However, the reaction times showed a larger reduction from A1 to A2 in the EG (85.90 % of the A1 value) compared to the CG (110.99 % of the A1 value). A one-tailed t-test for independent samples revealed a significant effect (t(31) = 2.23, p = .016, d=0.79, ΔM = 25.09, BCaCI₉₀% = [.81, 51.93]).

See Figure 2 and Table 1.

**Figure 2:**
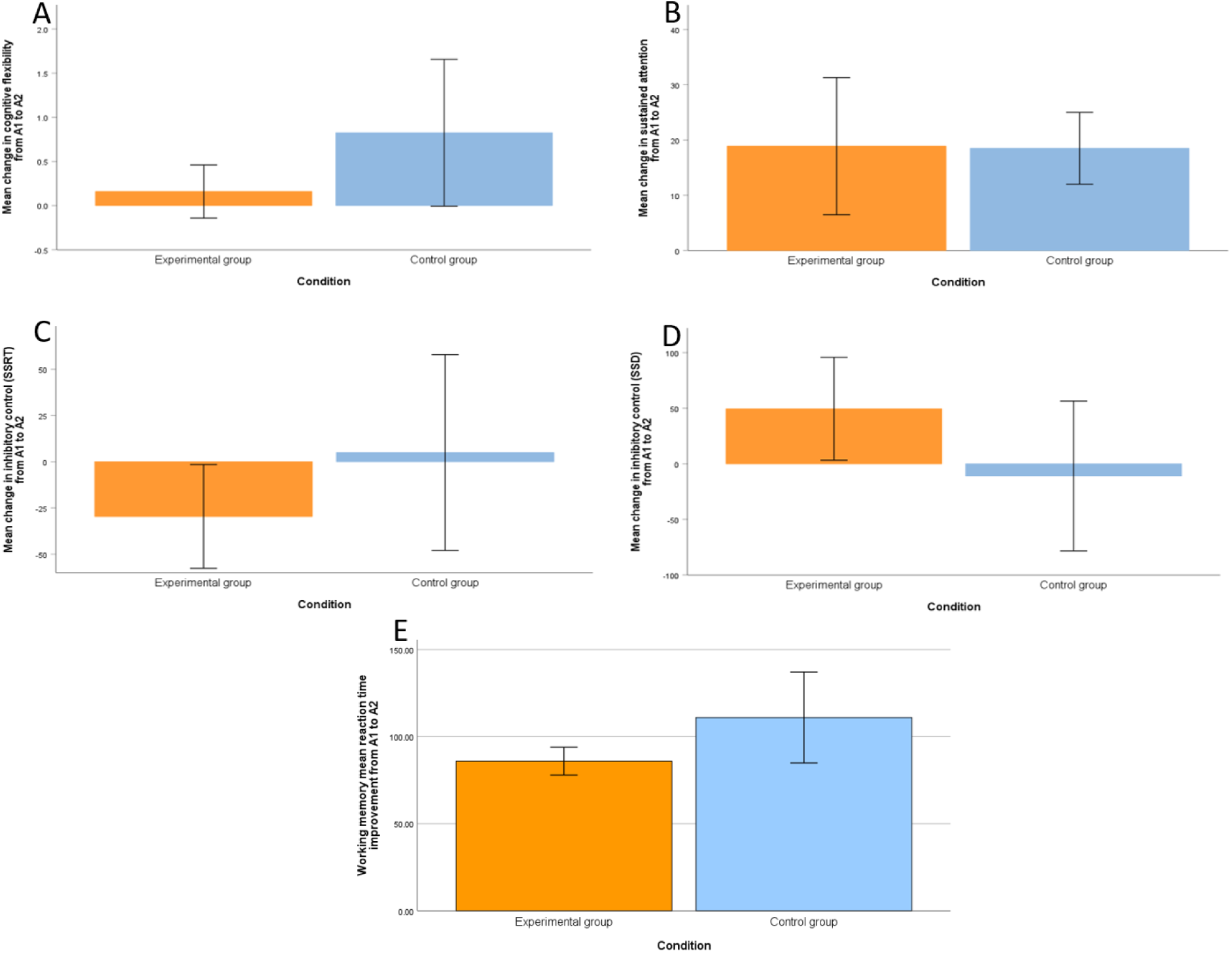
A: Mean change in cognitive flexibility from A1 to A2 for experimental and control group. B: Mean change in inhibitory control measured by the SSRT from A1 to A2 for experimental and control group. C: Mean change in inhibitory control measured by the SSD from A1 to A2 for experimental and control group. D: Mean change in sustained attention from A1 to A2 for experimental and control group. E: Mean reaction time improvement from A1 to A2 for experimental and control group. *Note*. The error bars represent the 90% confidence intervals.

**Table 1:**
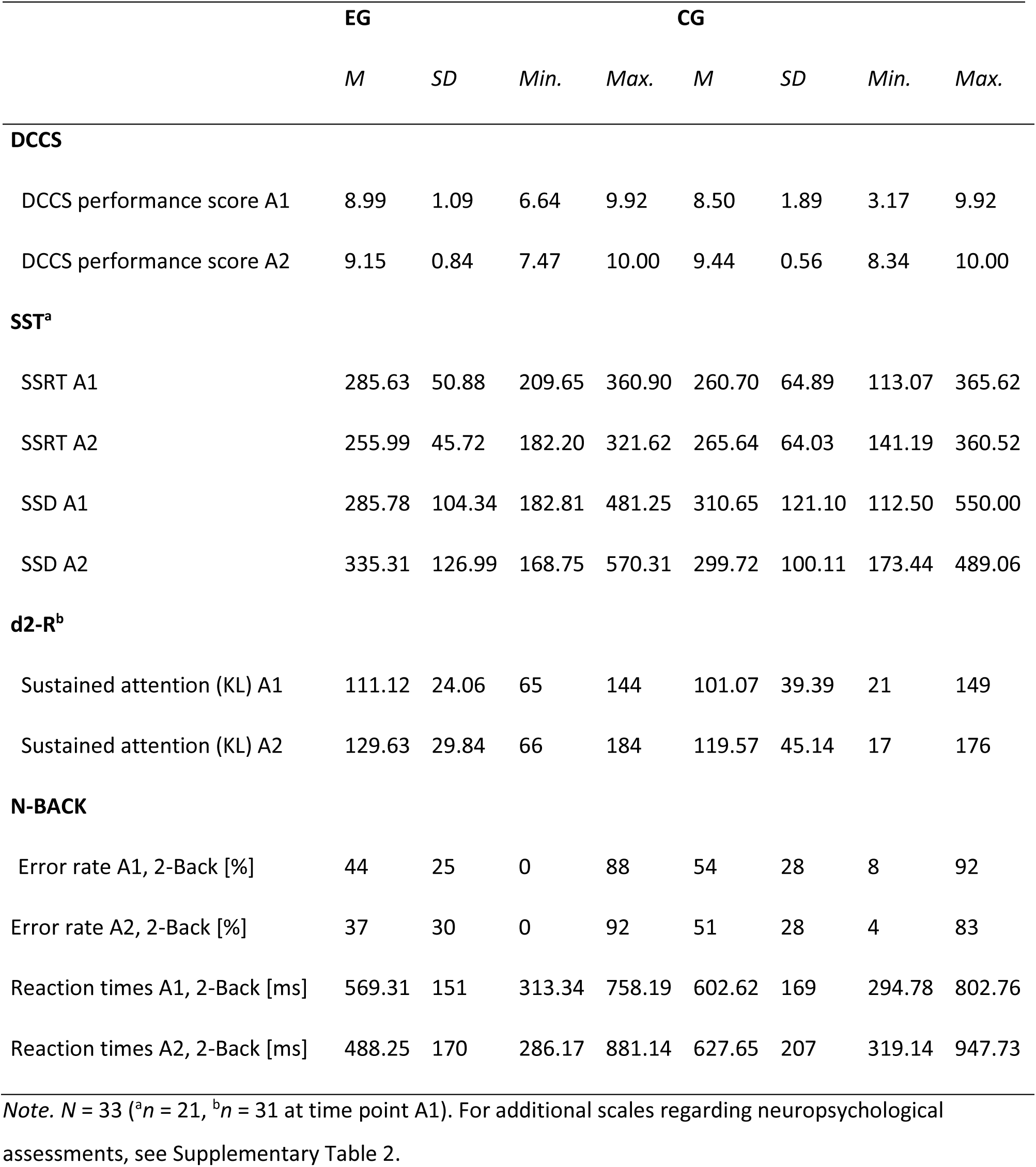
Descriptive data of neuropsychological assessments at time points A1 and A2 separated into experimental (EG) and control group (CG).

|  | **EG** |  |  |  | **CG** |  |  |  |
| --- | --- | --- | --- | --- | --- | --- | --- | --- |
|  | *M* | *SD* | *Min.* | *Max.* | *M* | *SD* | *Min.* | *Max.* |
| **DCCS** |  |  |  |  |  |  |  |  |
| DCCS performance score A1 | 8.99 | 1.09 | 6.64 | 9.92 | 8.50 | 1.89 | 3.17 | 9.92 |
| DCCS performance score A2 | 9.15 | 0.84 | 7.47 | 10.00 | 9.44 | 0.56 | 8.34 | 10.00 |
| **SST<sup>a</sup>** |  |  |  |  |  |  |  |  |
| SSRT A1 | 285.63 | 50.88 | 209.65 | 360.90 | 260.70 | 64.89 | 113.07 | 365.62 |
| SSRT A2 | 255.99 | 45.72 | 182.20 | 321.62 | 265.64 | 64.03 | 141.19 | 360.52 |
| SSD A1 | 285.78 | 104.34 | 182.81 | 481.25 | 310.65 | 121.10 | 112.50 | 550.00 |
| SSD A2 | 335.31 | 126.99 | 168.75 | 570.31 | 299.72 | 100.11 | 173.44 | 489.06 |
| **d2-R<sup>b</sup>** |  |  |  |  |  |  |  |  |
| Sustained attention (KL) A1 | 111.12 | 24.06 | 65 | 144 | 101.07 | 39.39 | 21 | 149 |
| Sustained attention (KL) A2 | 129.63 | 29.84 | 66 | 184 | 119.57 | 45.14 | 17 | 176 |
| **N-BACK** |  |  |  |  |  |  |  |  |
| Error rate A1, 2-Back [%] | 44 | 25 | 0 | 88 | 54 | 28 | 8 | 92 |
| Error rate A2, 2-Back [%] | 37 | 30 | 0 | 92 | 51 | 28 | 4 | 83 |
| Reaction times A1, 2-Back [ms] | 569.31 | 151 | 313.34 | 758.19 | 602.62 | 169 | 294.78 | 802.76 |
| Reaction times A2, 2-Back [ms] | 488.25 | 170 | 286.17 | 881.14 | 627.65 | 207 | 319.14 | 947.73 |

*Note.* $N = 33$ (<sup>a</sup> $n = 21$ , <sup>b</sup> $n = 31$ at time point A1). For additional scales regarding neuropsychological assessments, see Supplementary Table 2.

### Quality of Life

The EG showed a greater but non-significant improvement in physical well-being from A1 to A2 (M = 1.72, SD = 3.58) compared to the CG (M = 0.73, SD = 4.63), with a mean difference of ΔM = 0.99 (t(27) = 0.65, p = .261, d = 0.25, BCaCI₉₀% = [−1.57, 3.66]).

In terms of psychological well-being, the EG showed a significant medium improvement (M = 2.21, SD = 5.16) from A1 to A2, whereas the CG showed a decline (M = −1.46, SD = 5.85; group comparison t(30) = 1.87, p = .035, d = 0.67, BcaCI₉₀% = [0.38, 6.82]).

See Figure 3 and Table 2.

**Figure 3:**
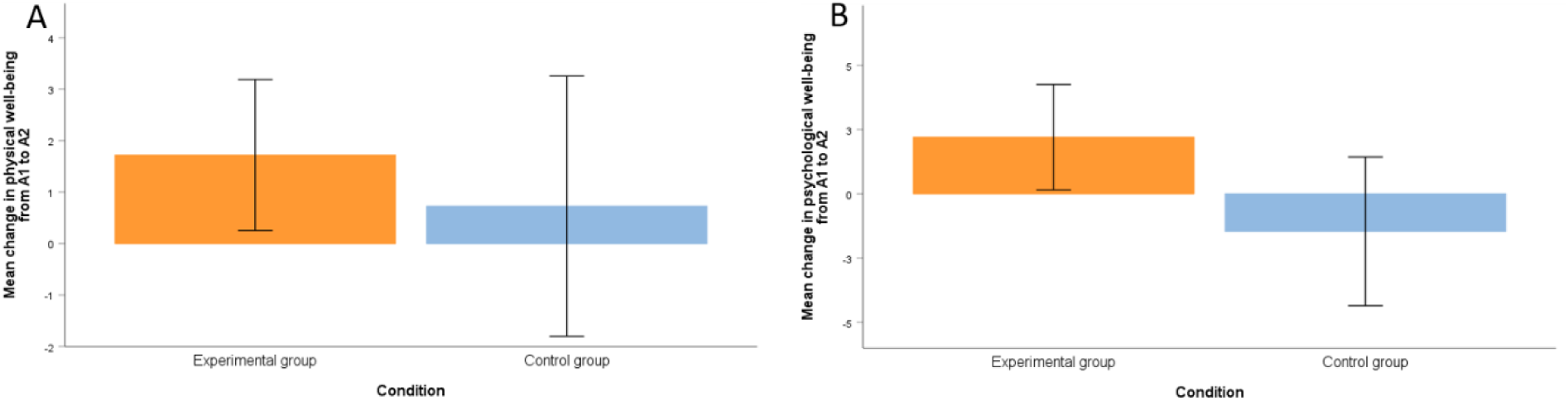
A: Mean change in physical well-being from A1 to A2 for experimental and control group. B: Mean change in psychological well-being from A1 to A2 for experimental and control group. *Note*. The error bars represent the 90% confidence intervals.

**Table 2:**
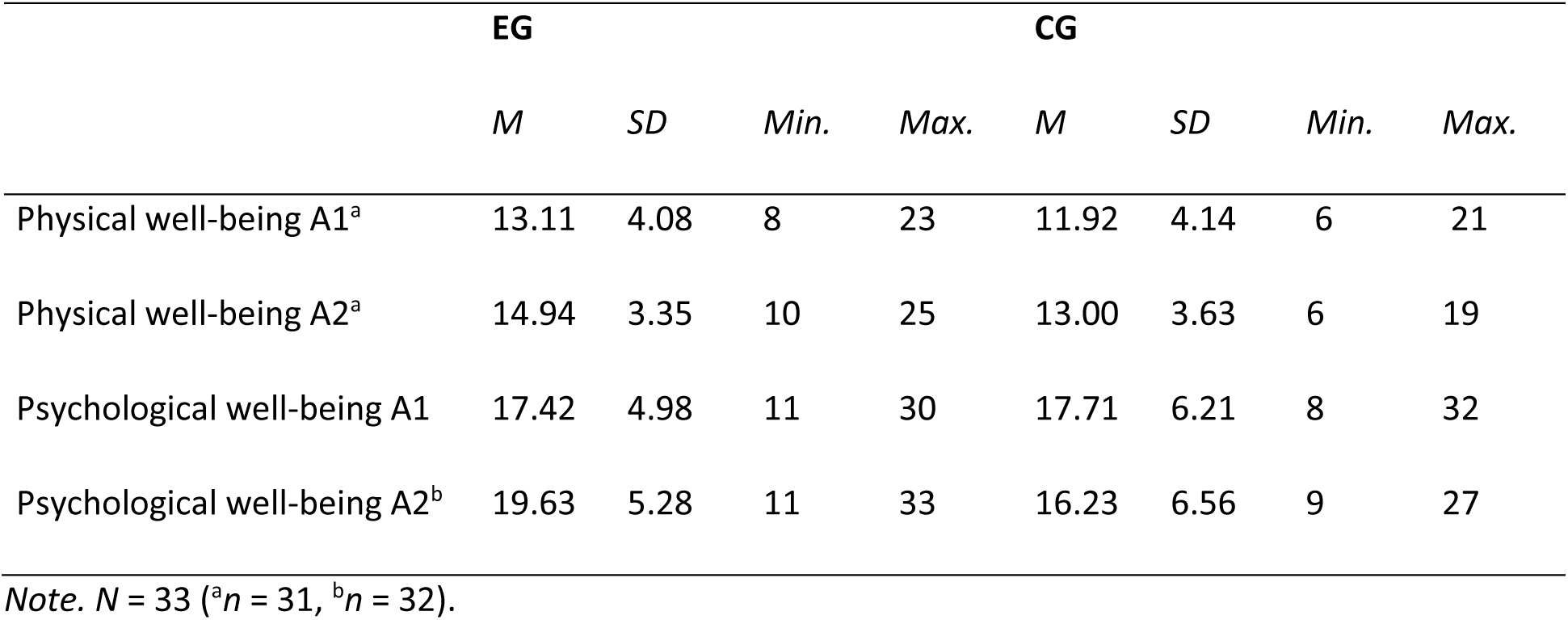
Descriptive data of physical and psychological well-being at time points A1 and A2 separated into experimental and control group.

|  | **EG** |  |  |  | **CG** |  |  |  |
| --- | --- | --- | --- | --- | --- | --- | --- | --- |
|  | *M* | *SD* | *Min.* | *Max.* | *M* | *SD* | *Min.* | *Max.* |
| Physical well-being A1<sup>a</sup> | 13.11 | 4.08 | 8 | 23 | 11.92 | 4.14 | 6 | 21 |
| Physical well-being A2<sup>a</sup> | 14.94 | 3.35 | 10 | 25 | 13.00 | 3.63 | 6 | 19 |
| Psychological well-being A1 | 17.42 | 4.98 | 11 | 30 | 17.71 | 6.21 | 8 | 32 |
| Psychological well-being A2<sup>b</sup> | 19.63 | 5.28 | 11 | 33 | 16.23 | 6.56 | 9 | 27 |

*Note.* *N* = 33 (<sup>a</sup>*n* = 31, <sup>b</sup>*n* = 32).

## DISCUSSION

The aim of this pilot study was to investigate whether a six-week chess training program, used as a therapeutic add-on, could improve cognitive functions and quality of life in adolescents with psychiatric disorders compared to treatment as usual (TAU) alone. A clinical quasi-experiment with pseudo-randomization was conducted, involving two measurement points, an experimental group, and a control group. The chess intervention followed the concept of “The King’s Plan for Kids”. While the hypotheses regarding improvements in cognitive flexibility, inhibitory control, sustained attention, and physical well-being were not confirmed, the data supported a significant improvement in psychological well-being and working memory efficiency following the six-week chess intervention.

Hypothesis 1 postulated that chess training would improve cognitive flexibility compared to TAU; however, this was not supported. In fact, results showed a slight trend in favor of the control group. Both groups improved over time, but many participants already scored highly at baseline, suggesting ceiling effects that limited potential gains. These findings, alongside small sample sizes, wide confidence intervals, and unequal variances, hinder definitive conclusions (Dumont et al., 2022; Eichorn et al., 2018; Lessing et al., 2019). Additionally, a pre-/self-selection bias may have resulted in cognitively stronger individuals opting for chess (Blanch, 2022). Although the EG may have employed more strategic thinking (Unterrainer et al., 2006), the results diverge from prior studies reporting positive effects of chess on cognitive flexibility (Goncalves et al., 2014; Grau-Pérez & Moreira, 2017; Ramos et al., 2018) and are more consistent with findings of limited or no effects (Sala & Gobet, 2016, 2017).

Hypothesis 2, which anticipated improved inhibitory control following chess training compared to TAU, was also not confirmed. However, the EG’s slower reaction times may indicate increased deliberation and cognitive control (Blasco-Fontecilla et al., 2016; ElDaou & El-Shamieh, 2015), despite the absence of statistical significance. The CG’s superior baseline performance suggests that the EG had greater potential for improvement.

Hypothesis 3 predicted that chess training would improve sustained attention, but this was not supported. Both groups showed improvement, with the EG starting from a higher baseline, potentially limiting observable gains. The broader range of maximum scores in the EG may reflect increased individual variability. This pattern could suggest a general training or practice effect— rather than a chess-specific benefit—as noted by Daseking and Putz (2015). These findings contrast with previous studies reporting benefits of chess for children with ADHD (Blasco-Fontecilla et al., 2016; ElDaou & El-Shamieh, 2015), healthy children (Aciego et al., 2012), or following cognitive training (Anagnostopoulou et al., 2019; Cortese et al., 2015).

An exploratory analysis of processing speed revealed small, non-significant effects in the EG, with a higher increase in processed target items—again diverging from prior findings. This inconsistent pattern underscores the challenge of interpretation, especially given the increased risk of Type I error across multiple tests (Bühner & Ziegler, 2017). Finally, non-significant results for accuracy (F%) should be interpreted cautiously, due to the measure’s limited reliability (Daseking & Putz, 2015) and potential fatigue effects, as the d2-R was administered last in the test battery.

In Hypothesis 4 we expected a beneficial effect of chess training on working memory. Although both groups did not show an increase in working memory performance (i.e. number of errors), the EG showed a larger improvement regarding faster reaction times from A1 to A2 compared to the CG while keeping the error rate stable. This could indicate an enhanced working memory efficiency in the EG which is suggested to be a beneficial effect of chess (review by Williams et al., 2025).

While the expected improvement in physical well-being (Hypothesis 5) was not confirmed, Hypothesis 6 was supported, with a significant improvement observed in psychological well-being. This aligns with the positive findings reported by Aciego et al. (2012) on the impact of chess on QoL in healthy children and adolescents—particularly in areas such as happiness, which can be considered indicators of QoL. It is also consistent with evidence from other forms of CRT in individuals with psychiatric disorders, both in adults (e.g., Tchanturia et al., 2017) and in children and adolescents (Pasqualotto et al., 2021).

The small, non-significant upward trend in physical well-being might be attributed to additional components of the intervention, such as physical activity (e.g., chess yoga). However, it remains unclear whether the observed effects stem directly from chess itself, the group-based social interaction, or simply the engagement in an additional structured activity alongside TAU. In contrast, any cognitive improvements observed are more likely to be directly linked to the chess training.

Overall, the findings suggest that the intervention holds promise for enhancing HRQoL in this population.

### Strengths and Limitations

A key strength of this study lies in its direct investigation of the effects of chess training in adolescents with psychiatric disorders—not only on cognitive functions but also on HRQoL, specifically physical and psychological well-being. While prior research has primarily focused on cognitive outcomes in typically developing children and adolescents, this study broadens the scope by including a clinical population. Notably, a significant improvement in psychological well-being was found, despite both groups receiving comprehensive TAU and the study’s small sample size.

However, several limitations may affect the generalizability of the findings. Mainly, the small and uneven sample likely influenced the results, as reflected in the wide confidence intervals. Replication with a larger and more balanced sample is necessary to confirm these findings and to detect smaller effects that may have gone unnoticed. Apparently, the intervention was not blinded, as participants naturally knew which group they were in, depending on whether they received training.

Pseudo-randomization may have introduced selection bias—participants in the chess group may have already been more cognitively proficient or intrinsically motivated (Blanch, 2022). Moreover, the absence of an active control group limits interpretability. A useful alternative might involve comparing the therapeutic chess intervention to classical chess instruction, given the broad framework of “The King’s Plan for Kids.”

Performance on cognitive tests may also have been affected by fatigue, particularly as the d2-R was administered last and without a break. Furthermore, information on participants’ prior chess experience or whether they continued playing chess beyond the intervention was not collected, which could influence skill transfer. Attendance varied, and the time between a participant’s final session and follow-up assessment was not documented—both factors that may impact outcomes. A promising avenue for future research is to explore how cognitive functioning relates to psychological well-being (e.g., Casey, 2023; Diamond, 2013), potentially as a mediating mechanism. For instance, it is plausible that improvements in executive functions contribute to better psychological outcomes.

### Clinical implications

This pilot study suggests that incorporating a chess training intervention into treatment as usual may offer clinically meaningful benefits for the psychological well-being of adolescents with psychiatric disorders. The medium-sized improvement in well-being, combined with high participant acceptance, highlights the potential of chess training as a low-risk, cost-effective, and easily implementable supplement to standard care. While effects on cognitive functioning were less robust—likely influenced by ceiling effects and the small sample size—the intervention may nonetheless promote greater reflection and more deliberate decision-making in some participants. Faster reaction times after chess training might suggest increased neural efficiency, which could be examined in future studies. Future research should seek to replicate these results in larger, more diverse samples, control for baseline differences, and further explore the mechanisms by which chess training may contribute to mental health and well-being in vulnerable youth.

## CONCLUSION

This pilot study provides valuable insights into the potential of chess as a therapeutic intervention for enhancing cognitive functioning and HRQoL in adolescents with psychiatric disorders. While the findings did not demonstrate significant effects on cognitive flexibility, inhibitory control, or sustained attention, they did indicate a positive impact on psychological well-being and potentially working memory. The medium-sized effect observed in the experimental group suggests that chess may serve as a promising supplementary intervention alongside TAU. Despite limitations such as the small sample size and challenges in assessing cognitive outcomes, the study highlights the feasibility and acceptance of chess training within a clinical population. Future research should involve larger samples, incorporate active control groups, and consider participants’ prior chess experience and continued engagement post-intervention. Overall, this study underscores the potential of chess as an accessible, engaging, and low-risk approach to supporting the mental health and well-being of adolescents facing psychiatric challenges.

## Authors Contribution

SH and AS recruited the participants and collected the data. AH, LW and KM helped with the recruitment of the participants and data collection. RS and AS conducted the chess training. SH, SG and SVK performed the data analysis. SG and SH wrote the manuscript. SVK and TB were responsible for the study design of the original study. SVK and TB procured the funding of the original study. All authors commented on the manuscript and provided intellectual input. All authors revised the manuscript critically for important intellectual content and approved the final version.

## Data Availability

The data that support the findings of this study are available on request from the corresponding author. The data are not publicly available due to privacy or ethical restrictions.

## Acknowledgments

We would like to thank all children and adolescence for their study participation. We further thank Paul Illner for his assistance with the ethics application and the study setup, Yury Shevchenko for programming the neuropsychological test battery, as well as Sophie Frahm and Ronja Bednar for her assistance in data collection. This project was supported in part by grants from the Münchener Schachstiftung (Munich Chess Foundation) and the Deutsche Forschungsgemeinschaft (German Research Foundation; Project ID 402170461, TRR 265). We further thank Grandmaster Stefan Kindermann for his support during the implementation of the chess training “Der Königsplan für Kinder”.

## Clinical trial registration

The original study was registered at www.clinicaltrials.gov (NCT01401491). Study procedures were in line with the recommendations of the World Medical Association (revised Declaration of Helsinki) as well as in accordance with good clinical practice guidelines, and were approved by the local Ethics Committee of the Medical Faculty Mannheim, Heidelberg University (approval # 2021-593).

