## Supplement for "Move by move towards mental health: A pilot study on chess as a therapeutic approach in adolescents with mental disorders"

Supplementary Table 1: Main discharge diagnoses of participants

|  | EG | | CG | | Full sample | |
| --- | --- | --- | --- | --- | --- | --- |
|  | *n* | % | *n* | % | *n* | % |
| F32.1 Major depressive disorder, single episode, moderate | 4 | 21.1 | 6 | 42.9 | 10 | 30.3 |
| F32.2 Major depressive disorder, severe single episode | 0 | 0.0 | 2 | 14.3 | 2 | 6.1 |
| F33.1 Major depressive disorder, moderate recurrent episode | 1 | 5.3 | 0 | 0.0 | 1 | 3.0 |
| F50.00 Anorexia nervosa, restrictive type | 4 | 21.1 | 2 | 14.3 | 6 | 18.2 |
| F50.1 Atypical anorexia nervosa | 1 | 5.3 | 0 | 0.0 | 1 | 3.0 |
| F50.2 Bulimia nervosa | 1 | 5.3 | 0 | 0.0 | 1 | 3.0 |
| F91.2 Conduct disorder, adolescent-onset type | 1 | 5.3 | 1 | 7.1 | 2 | 6.1 |
| F92.0 Depressive conduct disorder | 0 | 0.0 | 1 | 7.1 | 1 | 3.0 |
| F42.2 Mixed obsessional thoughts and acts | 0 | 0.0 | 1 | 7.1 | 1 | 3.0 |
| F40.1 Social phobias | 3 | 15.8 | 0 | 0.0 | 3 | 9.1 |
| F90.0 Attention-deficit hyperactivity disorder (ADHD) | 1 | 5.3 | 0 | 0.0 | 1 | 3.0 |
| F41.1 Generalized anxiety disorder | 1 | 5.3 | 0 | 0.0 | 1 | 3.0 |
| F41.2 Mixed anxiety and depressive disorder | 1 | 5.3 | 0 | 0.0 | 1 | 3.0 |
| F43.2 Adjustment disorder | 1 | 5.3 | 0 | 0.0 | 1 | 3.0 |
| F20.0 Paranoid schizophrenia | 0 | 0.0 | 1 | 7.1 | 1 | 3.0 |

*Note.* Classification according to ICD-10 (WHO, 2016)

Supplementary Table 2: Results of two-tailed independent samples t-tests assessing group comparability at A1 before the intervention for interval-scaled dependent and control variables

|  | Δ*M* | *t*(31) | *p* | Cohen’s *d* | BcaCI_95%_ |
| --- | --- | --- | --- | --- | --- |
| Age | -0.20 | -0.43^a^ | .675 | -0.16 | [-1.14, 0.74] |
| DCCS performance | 0.50 | 0.95 | .352 | 0.33 | [-0.44, 1.59] |
| SSRT | 24.93 | .97^b^ | .343 | 0.43 | [-21.07, 72.57] |
| SSD | -24.87 | -.50^b^ | .622 | -0.22 | [-117.69, 69.08] |
| d2-R KL | 10.05 | 0.87^c^ | .389 | 0.32 | [-11.83, 32.44] |
| Error rate A1, 2-Back [%] | -10.31 | -1.12 | .273 | -0.39 | [-28.42, 6.91] |
| Reaction times A1, 2-Back [ms] | -33.31 | -0.60 | .556 | -0.21 | [-140.87, 82.08] |
| PHY | 1.19 | 0.79^c^ | .439 | 0.29 | [-1.95, 4.28] |
| PWB | -0.29 | -0.15 | .881 | -0.05 | [-4.53, 3.69] |

*Note.* ^a^*t*(20.20), ^b^*t*(19), ^c^*t*(29)

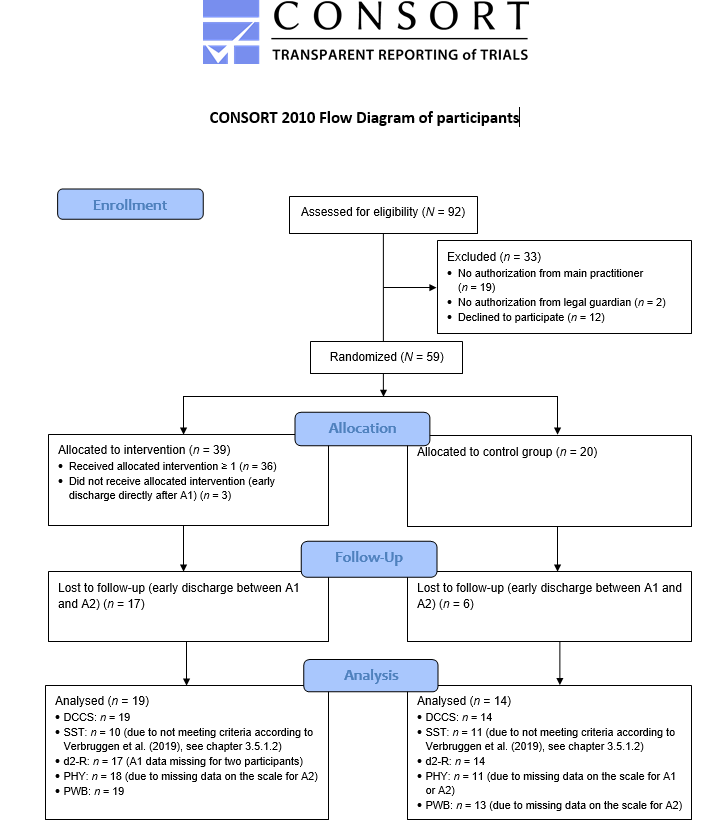

Supplementary Figure 1: CONSORT-Flow chart.
